# Novel associations of *BST1* and *LAMP3* with rapid eye movement sleep behavior disorder

**DOI:** 10.1101/2020.06.27.20140350

**Authors:** Kheireddin Mufti, Eric Yu, Uladzislau Rudakou, Lynne Krohn, Jennifer A. Ruskey, Farnaz Asayesh, Sandra B. Laurent, Dan Spiegelman, Isabelle Arnulf, Michele T.M. Hu, Jacques Y. Montplaisir, Jean-François Gagnon, Alex Desautels, Yves Dauvilliers, Gian Luigi Gigli, Mariarosaria Valente, Francesco Janes, Andrea Bernardini, Birgit Högl, Ambra Stefani, Evi Holzknecht, Karel Sonka, David Kemlink, Wolfgang Oertel, Annette Janzen, Giuseppe Plazzi, Elena Antelmi, Michela Figorilli, Monica Puligheddu, Brit Mollenhauer, Claudia Trenkwalder, Friederike Sixel-Döring, Valérie Cochen De Cock, Christelle Charley Monaca, Anna Heidbreder, Luigi Ferini-Strambi, Femke Dijkstra, Mineke Viaene, Beatriz Abril, Bradley F. Boeve, Jean-François Trempe, Guy A. Rouleau, Ronald B. Postuma, Ziv Gan-Or

## Abstract

**Objective:** To examine the role of genes identified through genome-wide association studies (GWASs) of Parkinson disease (PD) in the risk of isolated rapid-eye-movement (REM) sleep behavior disorder (iRBD).

**Methods:** We fully sequenced 25 genes previously identified in GWASs of PD, in a total of 1,039 iRBD patients and 1,852 controls. The role of rare heterozygous variants in these genes was examined using burden tests. The contribution of biallelic variants was further tested. To examine the potential impact of rare nonsynonymous *BST1* variants on the protein structure, we performed *in silico* structural analysis. Finally, we examined the association of common variants using logistic regression adjusted for age and sex.

**Results:** We found an association between rare heterozygous nonsynonymous variants in *BST1* and iRBD (*p*=0.0003 at coverage >50X and 0.0004 at >30X), mainly driven by three nonsynonymous variants (p.V85M, p.I101V and p.V272M) found in 22 (1.2%) controls vs. two (0.2%) patients. All three variants seem to be loss-of-function variants with a potential effect on the protein structure and stability. Rare non-coding heterozygous variants in *LAMP3* were also associated with iRBD (*p*=0.0006 at >30X). We found no association between rare heterozygous variants in the rest of genes and iRBD. Several carriers of biallelic variants were identified, yet there was no overrepresentation in iRBD.

**Conclusion:** Our results suggest that rare coding variants in *BST1* and rare non-coding variants in *LAMP3* are associated with iRBD. Additional studies are required to replicate these results and examine whether loss-of-function of *BST1* could be a therapeutic target.

## Introduction

Isolated rapid-eye-movement sleep behavior disorder (iRBD) is a prodromal synucleinopathy, as more than 80% of iRBD patients will eventually convert to Parkinson disease (PD, about 40-50% of patients), dementia with Lewy bodies (DLB) or unspecified dementia (40-50%), or, in much fewer cases, to multiple system atrophy (MSA, 5-10%).^1, 2^ While our understanding of the genetic background of DLB or MSA is limited, there are 80 genetic loci found to be associated with PD risk discovered through genome-wide association studies (GWASs),^3, 4^ and several genes have been implicated in familial PD.^5-7^

Recent studies have suggested that there is some overlap between the genetic backgrounds of iRBD and PD or DLB, yet this overlap is only partial. *GBA* variants are associated with iRBD risk, PD and DLB,^5, 8^ but pathogenic *LRRK2* variants are only associated with PD, and not with iRBD and DLB.^7, 9, 10^ *MAPT* and *APOE* haplotypes are important risk factors of PD and DLB, respectively,^11, 12^ but neither are linked to iRBD.^11, 13^ In the *SNCA* locus, specific variants in the 3’ untranslated region (UTR) are associated with PD but not with iRBD, and other, independent variants at 5’ UTR are associated with PD, iRBD and DLB.^14^

In the current study, we aimed to examine whether rare and common variants in 25 PD-related GWAS genes are associated with iRBD. Coding regions, exon-intron boundaries and 3’ and 5’ UTRs were fully captured and sequenced. We then performed different genetic analyses to investigate the association of these genes with iRBD.

## Materials and methods

### Study population

This study included a total of 2,891 subjects: 1,039 unrelated individuals who were diagnosed with iRBD (based on the International Classification of Sleep Disorders criteria, version 2 or 3) and 1,852 controls, all of European ancestry. The iRBD patients and controls were recruited through 18 centers in Canada and Europe, including centers from France, Germany, Austria, UK, Czech Republic, Italy, and Belgium. Diagnosis was performed with video polysomnography, the gold standard for iRBD diagnosis, by sleep specialists and neurologists. At recruitment, none of the patients had an overt neurodegenerative disease (PD, DLB, MSA etc.), which defined them as iRBD. In the current study, we had follow-up data available for 540 iRBD patients, 190 of which (35%) had converted since recruitment to PD, DLB or MSA or other overt neurodegenerative disease. The controls were not examined by polysomnography, but since only 1% of them is expected to develop iRBD, it is unlikely that this can affect the results. The European ethnicity of the participants was ascertained using GWAS data available in our lab, by performing principle component analysis and clustering with the European cohort available in HapMap v3. Details on age and on sex of the participants have been previously described,^15^ and data available from Dryad (Table e-1): https://doi.org/10.5061/dryad.vt4b8gtqd. Differences in age and sex were taken into account as needed in the statistical analysis. All patients and controls were of European ancestry (principal component analysis [PCA] of GWAS data was used to confirm ancestry, as compared to data from HapMap v.3 and hg19/GRCh37).

### Standard protocol approvals, registrations, and patient consents

The study participants signed informed consent forms at enrollment to the study, and the institutional review boards have approved the study protocol.

### Selection of genes and genetic analysis

We designed and performed the study prior to the publication of the recent PD GWAS,^3^ therefore, the genes for analysis were selected from previous GWASs.^16, 17^ A total of 25 genes were selected for analysis, including: *ACMSD, BST1, CCDC62, DDRGK1, DGKQ, FGF20, GAK, GPNMB, HIP1R, ITGA8, LAMP3, MAPT, MCCC1, PM20D1, RAB25, RAB29, RIT2, SETD1A, SLC41A1, STK39, SIPA1L2, STX1B, SYT11, TMEM163* and *USP25*. We selected the genes due to at least one of the following reasons: the gene has a quantitative trait loci association, the gene is expressed in the human brain, the gene potentially interact with PD-associated genes, or the gene is involved in pathways implicated in PD, for example the autophagy-lysosomal pathway, mitochondria quality control and endolysosomal recycling. The 25 genes were fully captured (coding sequence and 3’- and 5’-untranslated regions) as reported previously with molecular inversion probes (MIPs).^18^ The protocol is available on https://github.com/gan-orlab/MIP_protocol. Table e-2 details the probes used in the current study for the MIPs capture, data available from Dryad (Table e-2). Following the capture, we performed next-generation sequencing with the Illumina HiSeq 2500\4000 platform at the Génome Québec Innovation Centre. Sequencing reads were aligned to the hg19 reference genome using the Burrows-Wheeler Aligner.^19^ We then used the Genome Analysis Toolkit (GATK, v3.8) for quality control and to call variants,^20^ and ANNOVAR was used for variant annotation.^21^ We extracted the variant frequencies for the detected variant from the Genome Aggregation Database (GnomAD).^22^

### Quality control

To perform quality control (QC), we used PLINK v1.9.^23^ We excluded variants with a significant deviation (threshold set at *p*=0.001) from Hardy-Weinberg equilibrium among controls. Variants with <25% of their reads with a variant call were also excluded. To filter out variants with low genotyping rate, we set the threshold for inclusion on >90%. This threshold was also used to exclude samples from individuals with low call rates. when missingness rate was different among patients and controls (at *p*<0.05), we excluded these variants as well. variants with genotype quality score of <30 were further excluded. Rare variants (MAF 1%) were included based on two coverage thresholds, >30X and >50X, and all analyses were repeated using these thresholds. For the analysis of common variants, coverage of >15X was used.

### Statistical analysis

To test whether rare heterozygous variants (defined by MAF<0.01) in each of our target genes are associated with iRBD, we performed sequence kernel association test (SKAT, R package)^24^ and optimized SKAT (SKAT-O) on different groups of variants which included: all rare variants with MAF <1%, variants which are potentially functional (including splicing, nonsynonymous, stop-gain and frame-shift variants), only loss-of-function variants (stop-gain, frame-shift and splicing), and only nonsynonymous variants. In addition, we further used the Combined Annotation Dependent Depletion (CADD) score to test whether rare variants that are predicted to be pathogenic (based on a threshold of ≥12.37 which represents 2% of the variants predicted to be the most deleterious) are enriched in iRBD patients. To test the association between biallelic variants and iRBD risk, we used a threshold of MAF <0.1%, and examined whether there is enrichment of patients who carry two such variants compared to controls, using Fisher’s exact test. Variants included in this analysis were either splice-site, nonsynonymous, stop-gain or frame-shift variants. To properly account for multiple comparisons, in each analysis we applied Bonferroni correction that took into consideration the number of genes that were tested, and that the tests were done in two different depths of coverage, >30X and >50X. Therefore, when examining the association of 25 genes in two different depths of coverage, the threshold for statistical significance in this case was set on *p* = 0.05/50 = 0.001. To test for association of common variants (MAF>0.01) we used logistic regression adjusting for sex and age, using PLINK v1.9. Linkage disequilibrium between the variants we found and the respective GWAS top hits was examined using the reference cohort of non-Finnish European embedded in LDlink (https://ldlink.nci.nih.gov/).^25^ We used the genotype-tissue expression database (GTEx – https://www.gtexportal.org) to examine the effects of common variants on gene expression. We further performed in silico structural analysis of *BST1* to test whether the rare coding variants that were found to be associated with iRBD in our analysis could potentially affect the enzyme structure and activity. The atomic coordinates of human *BST1* bound to ATP-γ-S were downloaded from the Protein Data Bank (ID 1isg). We evaluated the steric clashes caused by the variants we identified using the “mutagenesis” toolbox in PyMol v. 2.2.0.

### Availability of data and materials

Data after processing that was used for the analyses in the current study are found in the supplementary tables, data available from Dryad (Table e-1 - e-7): https://doi.org/10.5061/dryad.vt4b8gtqd. The raw data can be requested from the corresponding author and will be shared anonymized.

## Results

### Coverage and identified variants

The average coverage of the 25 genes analyzed in this study was >647X (range 73-1162, median 790). An average of 95% of the target regions were covered with >15X, 93% with >30X and 90% with >50X. The average coverage of each gene and the percentage of the nucleotides covered at 15X, 30X and 50X are detailed in Table e-3, data available from Dryad (Table e-3). Finally, there were no differences in the coverage between patients and controls. A total of 1,189 rare variants were found with coverage of > 30X, and 570 rare variants with > 50X (data available from Dryad, Table e-4). We identified 125 common variants across all genes (data available from Dryad, Table e-5) with a coverage of >15X.

### Rare heterozygous variants in *BST1* and *LAMP3* are associated with iRBD

To examine whether rare heterozygous variants in our genes of interest may be associated with iRBD risk, we performed SKAT and SKAT-O tests, repeated twice for variants detected at depths of coverage of >30X and >50X (see methods). Table e-4 details all rare heterozygous variants identified in each gene and included in the analysis (data available from Dryad, Table e-4). We applied both SKAT and SKAT-O on five different groups of variants: all rare variants, all potentially functional variants (nonsynonymous, splice site, frame-shift and stop-gain), loss-of-function variants (frame-shift, stop-gain and splicing), nonsynonymous variants only, and variants with CADD score ≥12.37 (Table 1). The Bonferroni-corrected *p*-value threshold for statistical significance was set at *p*<0.001 after correcting for the number of genes and depths of coverage.

**Table 1.** Summary of results from burden analyses of rare heterozygous variants.

| DOC | Gene | All rare<br>(*p*-value) |  | Rare functional<br>(*p*-value) |  | Rare LOF<br>(*p*-value) |  | Rare NS<br>(*p*-value) |  | Rare CADD<br>(*p*-value) |  |
| --- | --- | --- | --- | --- | --- | --- | --- | --- | --- | --- | --- |
|  |  | SKAT-O | SKAT<br>Burden | SKAT-O | SKAT<br>Burden | SKAT-O | SKAT<br>Burden | SKAT-O | SKAT<br>Burden | SKAT-O | SKAT<br>Burden |
| 30x | *ACMSD* | 0.124 | 0.788 | 0.101 | 0.475 | NV | NV | 0.101 | 0.475 | 0.098 | 0.299 |
|  | *BSTI* | 0.040 | 0.020 | **0.0009** | **0.0004** | 0.348 | 0.653 | 0.001 | **0.0007** | 0.011 | 0.005 |
|  | *CCDC62* | 0.006 | 0.357 | 0.048 | 0.975 | 0.080 | 0.596 | 0.282 | 0.203 | 0.134 | 0.074 |
|  | *DDRGK1* | 0.152 | 0.112 | 0.745 | 0.527 | 0.765 | 0.587 | 0.746 | 0.742 | 0.919 | 0.677 |
|  | *DGKQ* | 0.067 | 0.062 | 0.067 | 0.062 | NV | NV | 0.183 | 0.188 | NV | NV |
|  | *FGF20* | 0.059 | 0.020 | 0.142 | 0.552 | 0.162 | 0.149 | 0.162 | 0.149 | 0.052 | 0.041 |
|  | *GAK* | 0.195 | 0.836 | 0.386 | 0.565 | 0.885 | 0.642 | 0.781 | 0.613 | 0.701 | 0.684 |
|  | *GPNMB* | 0.189 | 0.798 | 0.311 | 0.205 | 0.547 | 0.665 | 0.447 | 0.304 | 0.530 | 0.348 |
|  | *HIP1R* | 0.166 | 0.940 | 0.219 | 0.609 | NV | NV | 0.072 | 0.993 | 0.056 | 0.671 |
|  | *ITGA8* | 0.282 | 0.726 | 0.379 | 0.945 | 0.346 | 0.648 | 0.382 | 0.997 | 0.288 | 0.873 |
|  | *LAMP3* | **0.0006** | 0.478 | 0.189 | 0.322 | 0.787 | 0.300 | 0.318 | 0.724 | 0.524 | 0.601 |
|  | *MAPT* | 0.063 | 0.132 | 0.003 | 0.001 | 1 | 1 | 0.063 | 0.037 | 0.165 | 0.101 |
|  | *MCCC1* | 0.426 | 0.413 | 0.303 | 0.743 | 0.347 | 0.649 | 1 | 0.866 | 0.886 | 0.690 |
|  | *PM20D1* | 0.333 | 0.463 | 0.844 | 0.643 | 0.674 | 0.464 | 0.645 | 0.479 | 0.569 | 0.416 |
|  | *RAB25* | 0.561 | 0.395 | 0.807 | 0.802 | 0.777 | 0.968 | 0.565 | 0.637 | NV | NV |
|  | *RAB29* | 0.252 | 0.222 | 0.660 | 0.425 | NV | NV | 0.777 | 0.967 | 0.777 | 0.967 |
|  | *RIT2* | 0.395 | 0.242 | 0.023 | 0.266 | NV | NV | 0.576 | 0.495 | 0.576 | 0.495 |
|  | *SETD1A* | 0.667 | 0.500 | 0.241 | 0.452 | 0.171 | 0.166 | 0.300 | 0.940 | 0.310 | 0.399 |
|  | *SLC41A1* | 0.073 | 0.167 | 0.055 | 0.031 | 0.060 | 0.065 | 0.754 | 0.433 | 0.257 | 0.213 |
|  | *STK39* | 0.098 | 0.789 | 0.055 | 0.094 | 0.174 | 0.171 | 0.160 | 0.247 | 0.160 | 0.247 |
|  | *SIPA1L2* | 0.024 | 0.095 | 0.028 | 0.176 | 0.886 | 0.806 | 0.032 | 0.379 | 0.044 | 0.084 |
|  | *STX1B* | 0.504 | 0.682 | NV | NV | NV | NV | NV | NV | NV | NV |
|  | *SYT11* | 0.365 | 0.887 | 0.737 | 0.582 | 1 | 1 | 0.673 | 0.465 | 0.673 | 0.465 |
|  | *TMEM163* | 0.598 | 0.411 | 0.170 | 0.165 | NV | NV | 0.170 | 0.165 | 0.170 | 0.165 |
|  | *USP25* | 0.012 | 0.325 | 0.102 | 0.150 | NV | NV | 0.048 | 0.067 | 0.052 | 0.096 |
| 50x | *ACMSD* | 0.127 | 0.538 | 0.102 | 0.188 | NV | NV | 0.102 | 0.188 | 0.087 | 0.093 |
|  | *BSTI* | 0.018 | 0.009 | **0.0007** | **0.0003** | 0.671 | 0.470 | 0.001 | **0.0009** | 0.010 | 0.006 |
|  | *CCDC62* | 0.011 | 0.012 | 0.031 | 0.018 | 0.056 | 0.047 | 0.027 | 0.014 | 0.005 | 0.004 |
|  | *DDRGK1* | 0.368 | 0.314 | 0.899 | 0.811 | NV | NV | 0.899 | 0.811 | 0.899 | 0.811 |
|  | *DGKQ* | NV | NV | NV | NV | NV | NV | NV | NV | NV | NV |
|  | *FGF20* | 0.015 | 0.030 | 0.164 | 0.153 | 0.164 | 0.153 | NV | NV | 0.164 | 0.153 |
|  | *GAK* | 0.195 | 0.836 | 0.386 | 0.565 | 0.885 | 0.642 | 0.781 | 0.613 | 0.701 | 0.684 |
|  | *GPNMB* | 0.046 | 0.995 | 0.177 | 0.127 | 0.108 | 0.413 | 0.343 | 0.215 | 0.283 | 0.189 |
|  | *HIP1R* | NV | NV | NV | NV | NV | NV | NV | NV | NV | NV |
|  | *ITGA8* | 0.184 | 0.315 | 0.402 | 0.540 | 0.340 | 0.634 | 0.428 | 0.584 | 0.345 | 0.942 |
|  | *LAMP3* | 0.004 | 0.930 | 0.047 | 0.458 | 0.785 | 0.304 | 0.020 | 0.016 | 0.458 | 0.513 |
|  | *MAPT* | 0.605 | 0.503 | 0.619 | 0.659 | NV | NV | 0.619 | 0.659 | 0.619 | 0.659 |
|  | *MCCC1* | 0.26 | 0.151 | 0.972 | 0.752 | 0.334 | 0.618 | 0.951 | 0.745 | 0.958 | 0.800 |
|  | *PM20D1* | 0.903 | 0.934 | 0.873 | 0.758 | 0.662 | 0.487 | 1 | 0.758 | 0.816 | 0.666 |
|  | *RAB25* | 0.173 | 0.169 | NV | NV | NV | NV | NV | NV | NV | NV |
|  | *RAB29* | NV | NV | NV | NV | NV | NV | NV | NV | NV | NV |
|  | *RIT2* | 0.953 | 0.874 | 0.667 | 0.478 | NV | NV | 0.667 | 0.478 | 0.667 | 0.478 |
|  | *SETD1A* | 0.212 | 0.147 | 0.671 | 0.469 | NV | NV | 0.671 | 0.469 | 0.671 | 0.469 |
|  | *SLC41A1* | 0.132 | 0.189 | NV | NV | NV | NV | NV | NV | NV | NV |
|  | *STK39* | 0.073 | 0.808 | 0.051 | 0.087 | 0.160 | 0.146 | 0.160 | 0.146 | 0.160 | 0.146 |
|  | *SIPA1L2* | 0.163 | 0.098 | 0.598 | 0.861 | NV | NV | 0.777 | 0.963 | 0.777 | 0.963 |
|  | *STX1B* | NV | NV | NV | NV | NV | NV | NV | NV | NV | NV |
|  | *SYT11* | 0.0896 | 0.671 | 0.672 | 0.467 | NV | NV | NV | NV | NV | NV |
|  | *TMEM163* | 0.308 | 0.237 | 0.169 | 0.163 | NV | NV | 0.169 | 0.163 | 0.169 | 0.163 |
|  | *USP25* | 0.127 | 0.418 | 0.027 | 0.019 | NV | NV | 0.027 | 0.019 | 0.069 | 0.037 |

The table shows $p$ -values of the SKAT and SKAT-O analysis in the specified subgroups; in bold - result significant after Bonferroni correction for the number of genes and depths; DOC: Depth of coverage; LOF: Loss of function; NS: Nonsynonymous; CADD: Combined annotation dependent depletion; SKAT-O: Optimized sequence kernel association test; SKAT: Kernel association test; NV: No variants were found for this filter.

We found a statistically significant association of rare heterozygous functional variants in *BST1* (SKAT *p*=0.0004 at >30X and *p*=0.0003 at >50X for rare functional variants), found more in controls than in iRBD patients. This association is mainly driven by the nonsynonymous variants p.V85M (rs377310254, found in five controls and none in patients), p.I101V (rs6840615, found in seven controls and none in patients), and p.V272M (rs144197373, found in 10 controls and two patients). Overall, these variants were found in 22 (1.2%) controls vs. 2 (0.2%) patients. Of these two patients, one had self-reported age at onset of 25 years old (not confirmed by polysomnography at the time) and he was 75 years old at the time of the study, and did not convert to an overt synucleinopathy. The second patient had age at onset of 71 years old (compared to an average age at onset of 61 years in the entire cohort) and converted to DLB at the age of 73.

Another statistically significant association was found between rare variants in *LAMP3* gene and reduced iRBD risk in SKAT-O analysis. This association is driven by two non-coding variants (one intronic [location - chr3:182858302] and one at the 3’ UTR of *LAMP3* [rs56682988, c.*415T>C]) found only in controls (15 and nine controls, respectively). In order to further examine whether these variants indeed drive the association in both *BST1* and *LAMP3*, we excluded them and repeated the analysis (SKAT and SKAT-O), which resulted in loss of statistical significance for both genes (data available from Dryad, Table e-6). There were no additional statistically significant associations of the remaining genes with iRBD after correcting for multiple comparisons (*p*<0.001). We also repeated the analysis in 350 patients with available data on conversion who did not convert at the time of the study (data available from Dryad, Table e-7). Rare functional and rare nonsynonymous *BST1* variants were nominally associated with iRBD (*p*=0.021 and *p*=0.049, SKAT), but not after correction for multiple comparisons. The association of *LAMP3* had a *p*=0.06 (SKAT-O), demonstrating the reduced power in this smaller sub-population.

### Structural analysis of *BST1* variants suggests that loss-of-function may be protective in iRBD

To investigate the potential impact of the three *BST1* nonsynonymous variants (p.V85M, p.I101V and p.V272M) on the structure and activity of the enzyme, we performed *in silico* mutagenesis and evaluated potential clashes with surrounding residues. Figure 1 depicts the structure of *BST1* with the respective locations of the three nonsynonymous variants that drive the *BST1* association detected in our analysis. The structure of human *BST1* was solved by X-ray crystallography in complex with five substrate analogues ^26^. All structures revealed a homodimeric assembly, with the catalytic clefts facing the cavity at the interface of the two chains (Figure 1A).

**Figure 1.**
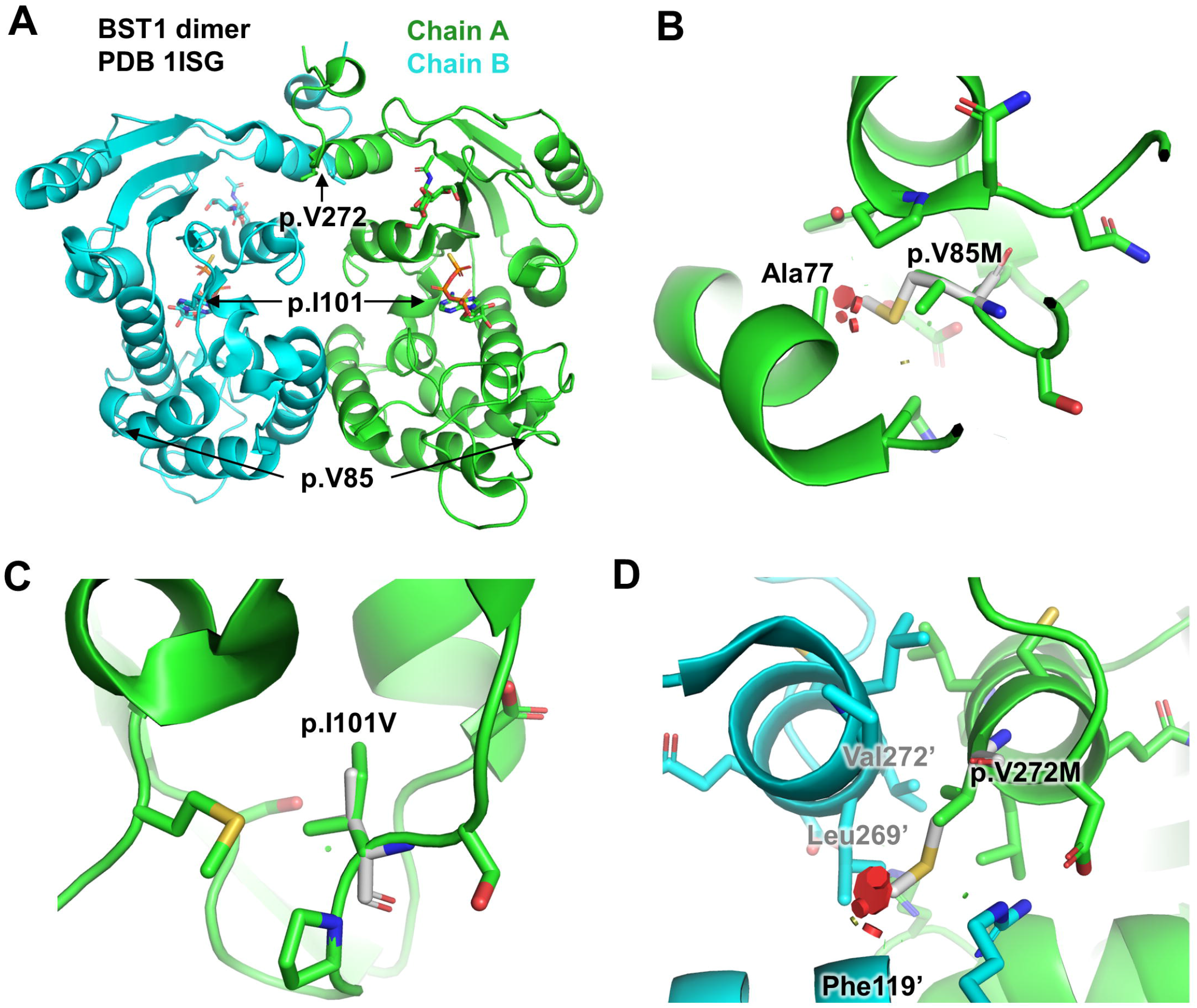
Structural analysis of human *BST1* variants. This figure was produced using the software PyMOL v.2.2.0, and represents: (A) Structure of the BST1 dimer bound to ATP-γS (pdb 1ISG). The position of each variant sites is indicated. The ATP-γS molecule in the active site is shown as sticks. (B) Close-up view of the p.V85M variant site. The mutated residue is shown in white. The variant would create clashes (red disks) with nearby Ala77 in the core. (C) Close-up view of the p.I101V variant site. The residue is located in the core of the protein, but the variant to a smaller residue results in no clash. (D) Close-up view of the p.V272M variant site. Primed (‘) residues correspond to chain B. This residue is located at the dimer interface and the variant would create clashes with the other chain resulting in a destabilization of the dimer.

The sidechain of p.V85M points towards the hydrophobic core of the protein, behind a helix facing the nucleotide binding site. The amino-acid change from valine to the bulkier sidechain of methionine results in clashes with other residues in the core, for all rotamers (Figure 1B). This variant would therefore likely destabilize the enzyme active site and potentially unfold the protein. The sidechain of the variant p.I101V is located underneath the active site towards the hydrophobic core. Although the amino-acid change from isoleucine to the smaller sidechain of valine does not create clash (Figure 1C), it reduces the packing in the core, which could also destabilize the enzyme. Finally, the p.V272M variant is located in a helix at the C-terminus of the protein that forms symmetrical contacts with the same helix in the other chain of the dimer. The p.V272M variant would create clashes with sidechain and main-chain atoms located in the other chain of the dimer (Figure 1D). As p.V272M resides at the dimer interface of the enzyme and probably helps maintaining the two subunits together, this variant would most likely lead to the disruption of the dimer. Overall, all the disease-associated nonsynonymous variants in *BST1* (p.V85M, p.I101V, and p.V272M) appear to be “loss-of-function”, suggesting that reduced BST1 activity may be protective in iRBD. This is supported by the top PD GWAS hit in the *BST1* locus, the rs4698412 G allele, which is associated with reduced risk of PD ^3^. This allele is also associated with reduced expression of BST1 in blood in GTEx (normalized effect size =-0.07, *p*=1.5e-6), suggesting that reduced expression might be protective.

### Very rare bi-allelic variants are not enriched in iRBD patients

In order to examine whether bi-allelic variants in our genes of interest are enriched in iRBD, we compared the carrier frequencies of very rare (MAF<0.001) homozygous and compound heterozygous variants between iRBD patients and controls. To analyze compound heterozygous variants, since phasing could not be performed, we considered carriers of two very rare variants as compound heterozygous carriers, with the following exceptions: 1) when variants were physically close (less than 112 base pairs [bp]; probes’ target length) and we could determine their phase based on the sequence reads, and 2) if the same combination of very rare variants appeared more than once across samples, we assumed that the variants are most likely to be on the same allele. We found five (0.5%) iRBD patients and seven (0.4%) controls presumably carriers of bi-allelic variants in the studied genes (Table 2, *p*=0.731, Fisher test).

**Table 2.**
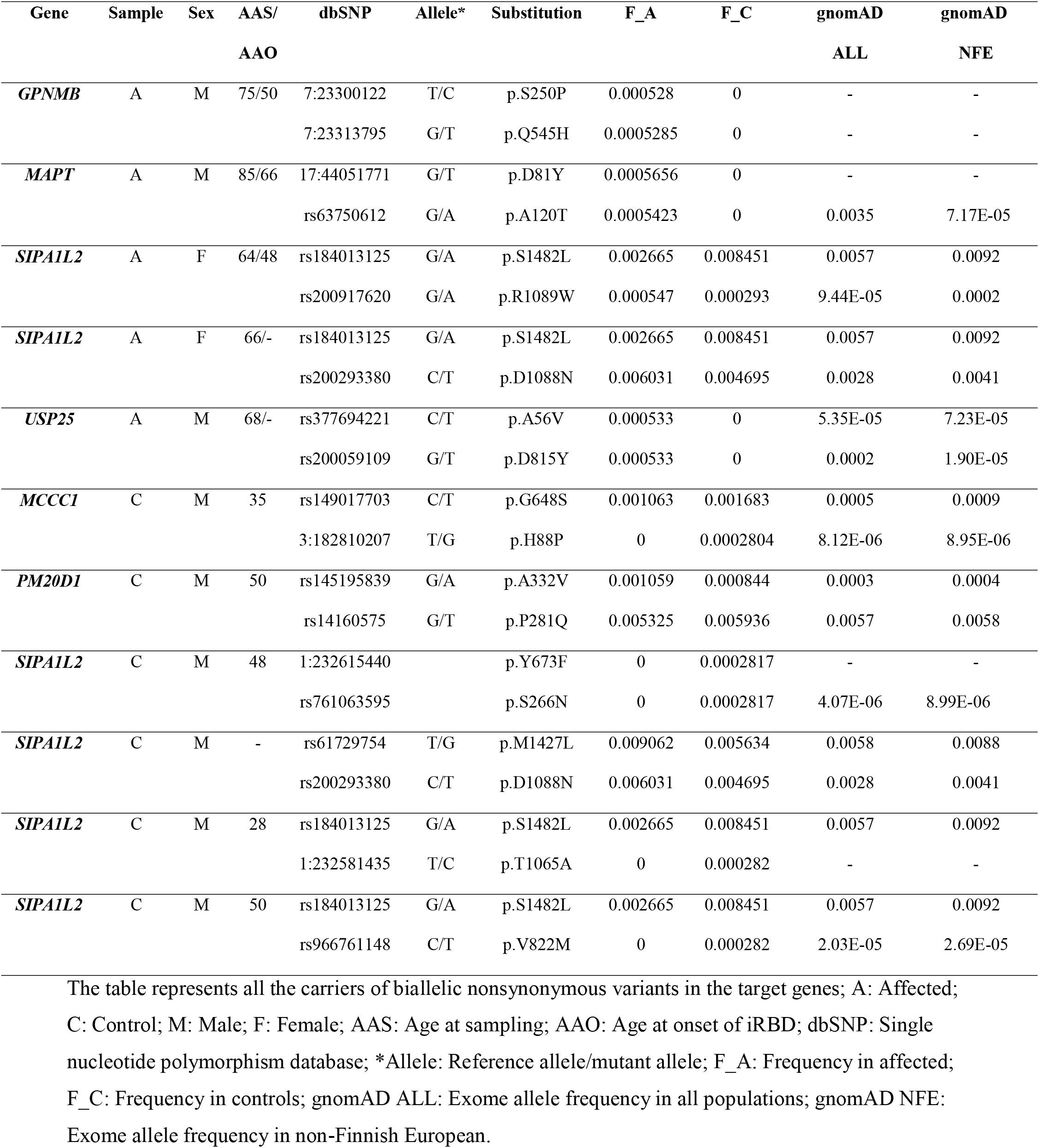
Summary of all samples carrying two nonsynonymous variants detected in the present study.

### Association of common variants in the target genes with iRBD

To test whether common variants in our target genes are associated with iRBD, we performed logistic regression (using PLINK v1.9 software) adjusted for age and sex for common variants (MAF>0.01) detected at coverage depth of >15X. A nominal association was observed in 12 variants across all genes (data available from Dryad, Table e-5), but no association remained statistically significant after Bonferroni correction for multiple comparisons (set at *p*<0.0005).

Of the variants with nominal associations, one variant in the *ITGA8* 3’ UTR (rs896435, OR=1.15, 95% CI = 1.01-1.32, *p*=0.04) is the top hit from the most recent PD GWAS,^3^ and two other *ITGA8* 3’ UTR variants are almost in perfect LD (D’=1.0, R^2^>0.99, *p*<0.0001) with rs896435. Four variants in the 3’ UTR of *RAB29* were almost in perfect LD (data available from Dryad, Table e-5) and are associated with expression of RAB29 in multiple tissues in GTEx, including the brain. Three *MAPT* variants were in partial LD with PD GWAS hits in the *MAPT* locus and were associated with expression of multiple genes in multiple tissues in GTEx, demonstrating the complexity of this genomic region.

## Discussion

In the current study, we studied a large cohort of iRBD patients by fully-sequencing and analyzing 25 PD-related GWAS genes and their association with iRBD. Our results identify *BST1* and *LAMP3* as novel genes potentially associated with iRBD. Based on *in silico* models, the three nonsynonymous *BST1* variants that drive the association with iRBD may be loss-of-function variants, suggesting that reduced *BST1* activity may reduce the risk of developing iRBD. This hypothesis is further supported the results from the PD GWAS, in which the *BST1* locus variants were associated with reduced risk of PD were also associated with reduced expression of *BST1*.^3^ Further studies will be required to determine that these variants are indeed loss-of-function variants and to study the mechanism underlying this potential protective association. The variants driving the association of *LAMP3* are in noncoding regions and could be regulatory. These hypotheses will require confirmation in functional studies in relevant models. While some common variants were nominally associated with iRBD, none of them remained statistically significant after correction for multiple comparisons.

This study provides further support to our previous studies, showing that there is only partial overlap between the genetics of iRBD and PD. Genes that are important in PD such as *LRRK2* and *MAPT* seem to have no role in iRBD,^10,11^ whereas other genes such as *GBA* are important in both, as well as in DLB.^5, 8^ However, we cannot rule out that some of the 25 genes that we tested in the current study are associated with iRBD, yet the effect size of the association is too small to detect with the current sample size. Therefore, additional studies in larger cohorts of iRBD will be required in the future to fully uncover the association between PD-related genes and iRBD.

BST1, also called CD157, is a glycosyl phosphatidylinositol (GPI) anchored membrane protein initially found in bone marrow stromal cells and is essential for B-lymphocyte growth and development. It has an extracellular enzymatic domain that produces cyclic ADP-ribose (cADPR). This metabolite acts as a second messenger that can trigger Ca^2+^ release from intracellular stores,^27^ a process that plays a role in cellular function and death. Specific features of calcium homeostasis have been suggested to be responsible for the specific vulnerability of dopaminergic neurons in PD,^28^ yet whether BST1 is involved in calcium homeostasis in human neurons is still unclear, as most work was done in non-human models. Another mechanism by which BST1 may be involved in PD is immune response and neuroinflammation, which are likely important in the pathogenesis of the disease. BST1 serves as a receptor which regulates leukocyte adhesion and migration, and plays a role in inflammation.^29^ However, its potential role in microglia activation and neuroinflammation is yet to be determined. Our *in-silico* analysis suggested that the *BST1* variants found mostly in controls are loss-of-function variants. Furthermore, common variants in the *BST1* locus, carried by >50% of the European population, are associated with reduced risk of PD in the most recent GWAS, and are also associated with reduced expression of *BST1* in blood,^3^ indicating that reduced *BST1* expression may be protective. We can therefore hypothesize that these variants may reduce immune response and lead to a reduced risk of iRBD, and that inhibition of *BST1* could become a target for future preclinical drug discovery efforts for iRBD and PD treatment or prevention. Understanding the role of *BST1* as an immune response gene in iRBD, understanding the mechanism underlying this association, and examining whether it can serve as a target for early therapeutics development, will require additional research in humans and different models.

*LAMP3* encodes the lysosomal-associated membrane protein 3, which plays a role in the unfolded protein response (UPR) that contributes to protein degradation and cell survival during proteasomal dysfunction.^30^ Furthermore, LAMP3 knockdown impairs the ability of the cells to complete the autophagic process, and high LAMP3 expression is associated with increased basal autophagy levels.^31^ Numerous PD-related genes have been implicated in the autophagy-lysosomal pathway,^32^ and genes associated with iRBD such as *GBA*,^*33*^ *TMEM175*^*34*^ and *SNCA*^*14*^ are all involved in this pathway. Our current findings further strengthen the potential association between the autophagy-lysosomal pathway and iRBD. The specific role of LAMP3 in this pathway and how it may be involved in iRBD and the subsequent synucleinopathies will require additional studies. Since the rare variants we identified were found only in controls, it may suggest that LAMP3 may have a protective effect that should be further explored.

Our study has several limitations. First, despite being the largest genetic study of iRBD to date, it may be still underpowered to detect rare variants in GWAS PD-related genes, as well as common variants with a small effect size. Therefore, we cannot completely rule out the possibility that rare and common variants in these genes may contribute to iRBD risk. A second limitation is the younger age and the differences in sex distribution between iRBD patients and controls, for which we adjusted in the statistical analysis as needed. Another potential limitation is the possibility that there were undiagnosed iRBD patients among the control population. However, since iRBD is found in only ∼1% of the population,^1^ the effect of having undiagnosed iRBD patients in the controls would be minimal, given the large sample size. Additional limitation is the lack of replication cohort; therefore, our findings should be replicated in additional iRBD cohorts once they become available. Lastly, our study includes only individuals of European descent, and fully understanding the genetics of iRBD will require studies with other ethnicities. Unfortunately, due to the reduced availability of polysomnography in many countries, there are currently only very few small cohorts of iRBD patients from other populations, and more effort is required to develop such cohorts in different countries.

To conclude, our results suggest two novel genetic associations with iRBD; an association with rare functional variants in *BST1*, and with rare non-coding variants in *LAMP3*. All the association-driving coding variants found in *BST1*, mainly in controls, appear to potentially cause loss-of-function, suggesting that reduced BST1 activity may reduce the risk of iRBD. Further studies would be required to confirm our results and to examine the biological mechanism underlying the effect of disease-associated variants in both *LAMP3* and *BST1*. The absence of evidence of association between rare and common variants in the remaining genes and iRBD risk suggests that these genes either have no effect in iRBD or have a minor effect that we could not detect with this sample size. Environmental factors and environment-gene interactions are likely to play a major role on iRBD, and larger studies that include carefully collected epidemiological data and more extensive genetic data such as whole-exome or whole-genome sequencing will be required to clarify these issues.

## Glossary

AAO: age at onset
AAS: age at sampling
*ACMSD*: Aminocarboxymuconate semialdehyde decarboxylase
*BST1*: Bone marrow stromal cell antigen 1
CADD: Combined Annotation Dependent Depletion
*CCDC62*: Coiled-coil domain containing 62
CI: confidence interval
DOC: depth of coverage
F_A: frequency in affected
F_C: frequency in control
*FGF20*: Fibroblast growth factor 20
*GAK*: Cyclin G associated kinase
GATK: Genome Analysis Toolkit
GnomAD: Genome Aggregation Database
*GPNMB*: Glycoprotein (transmembrane) nmb
GTEx: Genotype-tissue expression
GWAS: genome-wide association study
hg19: human genome version 19
*HIP1R*: Huntingtin interacting protein 1 related
iRBD: isolated rapid-eye-movement sleep behavior disorder
*ITGA8*: Integrin subunit alpha 8
*LAMP3*: Lysosomal-associated membrane protein 3
LD: linkage disequilibrium
LOF: loss-of-function
MAF: minor allele frequency
*MAPT*: Microtubule-associated protein tau
*MCCC1*: Methylcrotonoyl-CoA carboxylase 1 (α)
MIPs: molecular inversion probes
NS: non-synonymous
OR: odds ratio
PCA: principle component analysis
*PM20D1*: Peptidase M20 domain containing 1
QC: quality control
QS: quality score
REM: rapid eye movement
*RIT2*: Ras Like without CAAX 2
*SCARB2*: Scavenger receptor class B, member 2
*SETD1A*: SET domain containing 1A, histone lysine methyltransferase
*SIPA1L2*: Signal induced proliferation associated 1 like 2
SKAT: sequence kernel association test
SKAT-O: optimized SKAT
*SLC41A1*: Solute carrier family 41 (magnesium transporter), member 1
*STK39*: Serine/threonine kinase 39
*STX1B*: Syntaxin 1B
SYT11: Synaptotagmin 11
*TMEM163*: Transmembrane protein 163
*USP25*: Ubiquitin specific peptidase 25
UTR: untranslated region
vPSG: video polysomnography

## Acknowledgments

We thank the participants for contributing to this study. JYM holds a Canada Research Chair in Sleep Medicine. JFG holds a Canada Research Chair in Cognitive Decline in Pathological Aging. WO is Hertie Senior Research Professor, supported by the Hertie Foundation. EAF holds a Canada Research Chair (Tier 1) in Parkinson disease. GAR holds a Canada Research Chair in Genetics of the Nervous System and the Wilder Pen field Chair in Neurosciences. JFT holds a Canada Research Chair (Tier 2) in Structural Pharmacology. ZGO is supported by the Fonds de recherche du Québec–Santé Chercheur-Boursier award and is a Parkinson Canada New Investigator awardee. We thank D. Rochefort, H. Catoire, and V. Zaharieva for their assistance.

## Appendix 1 Authors

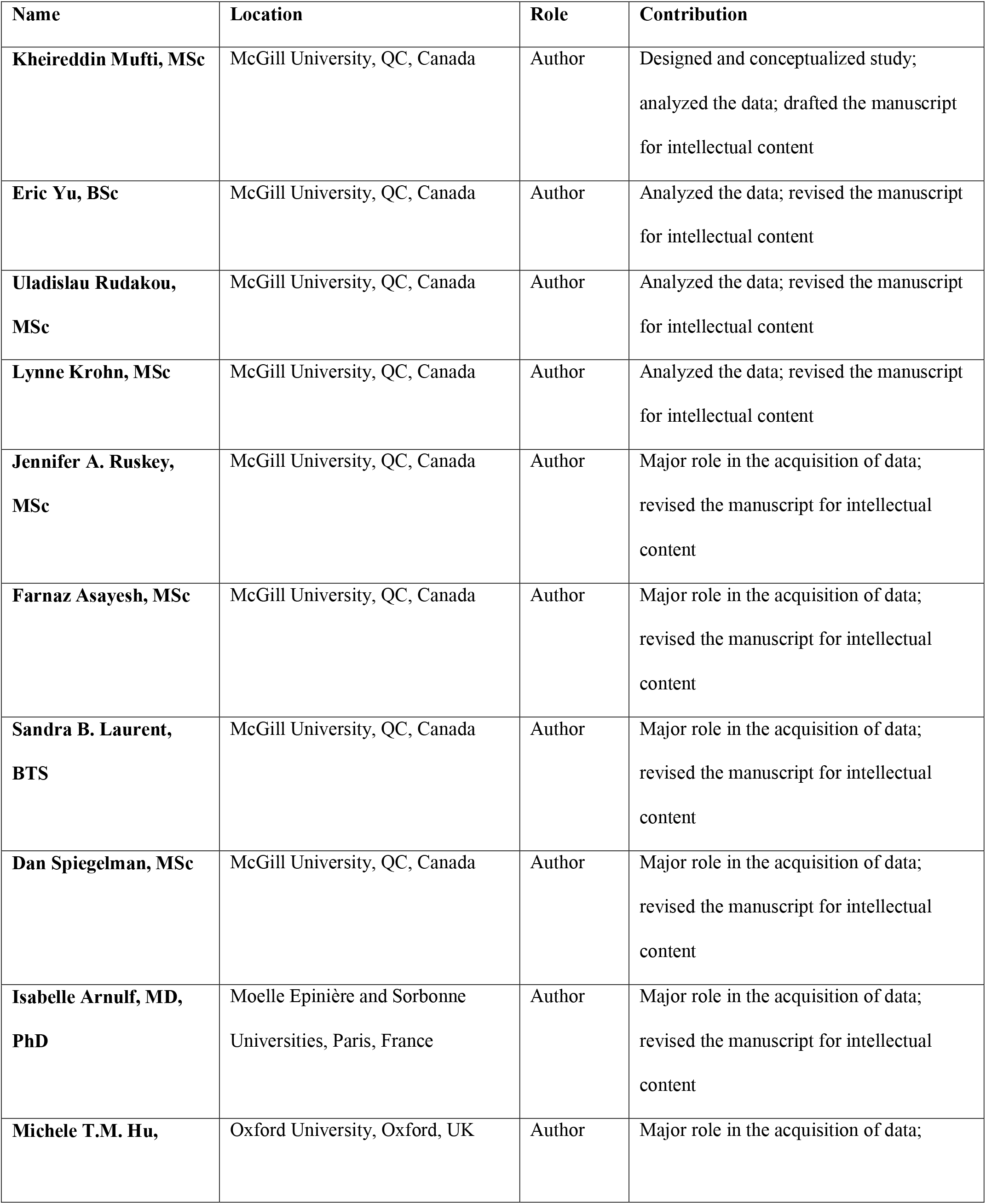
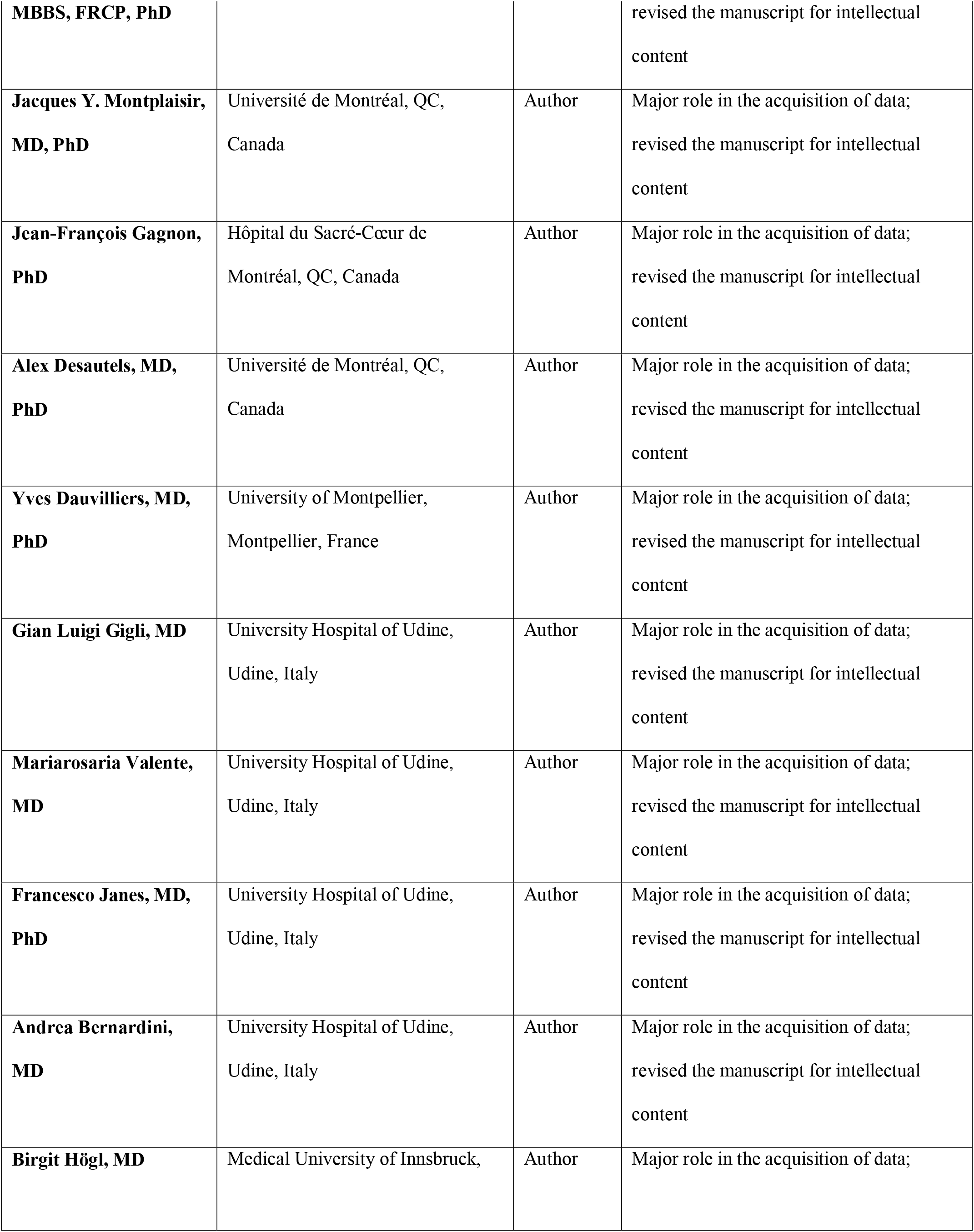
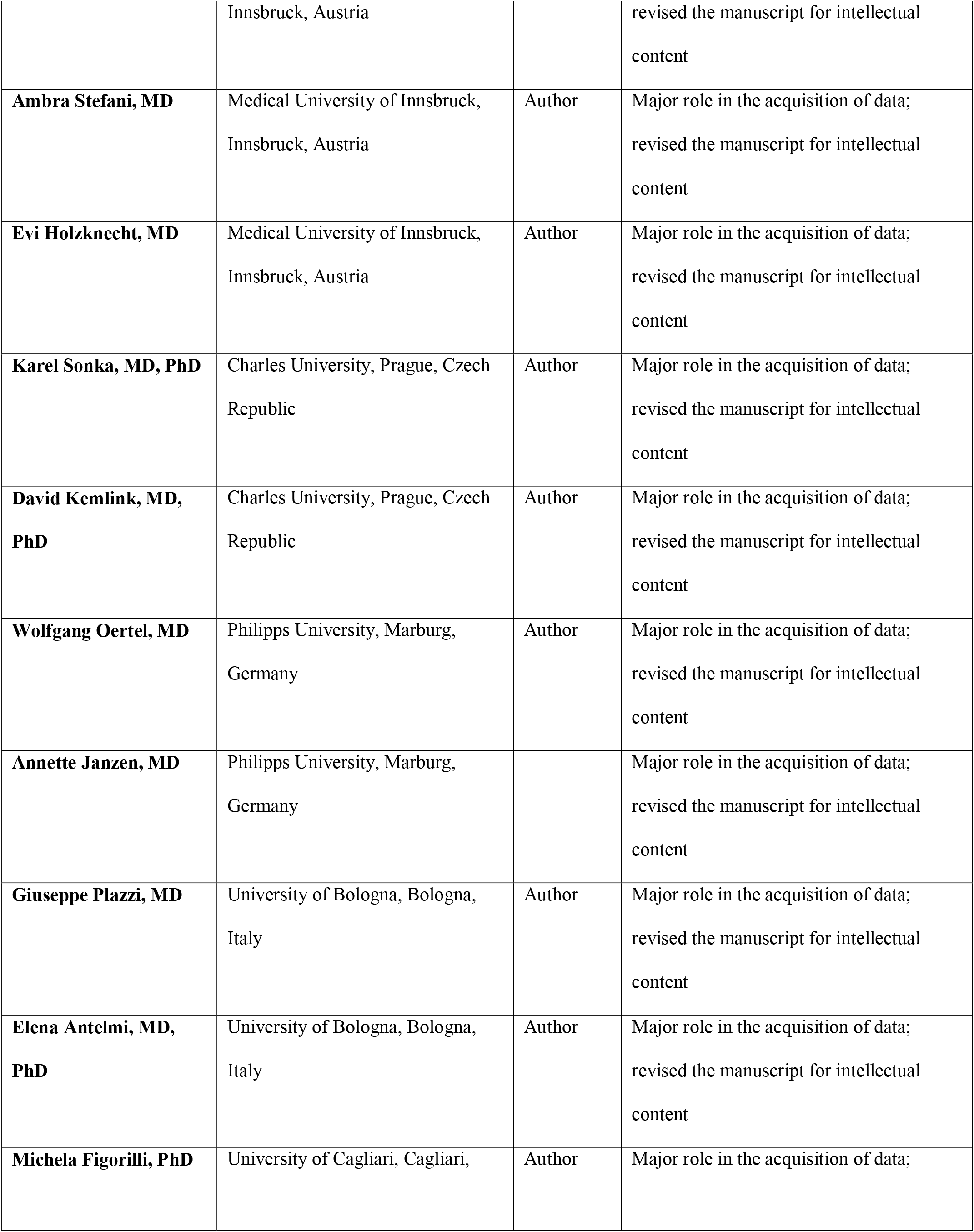
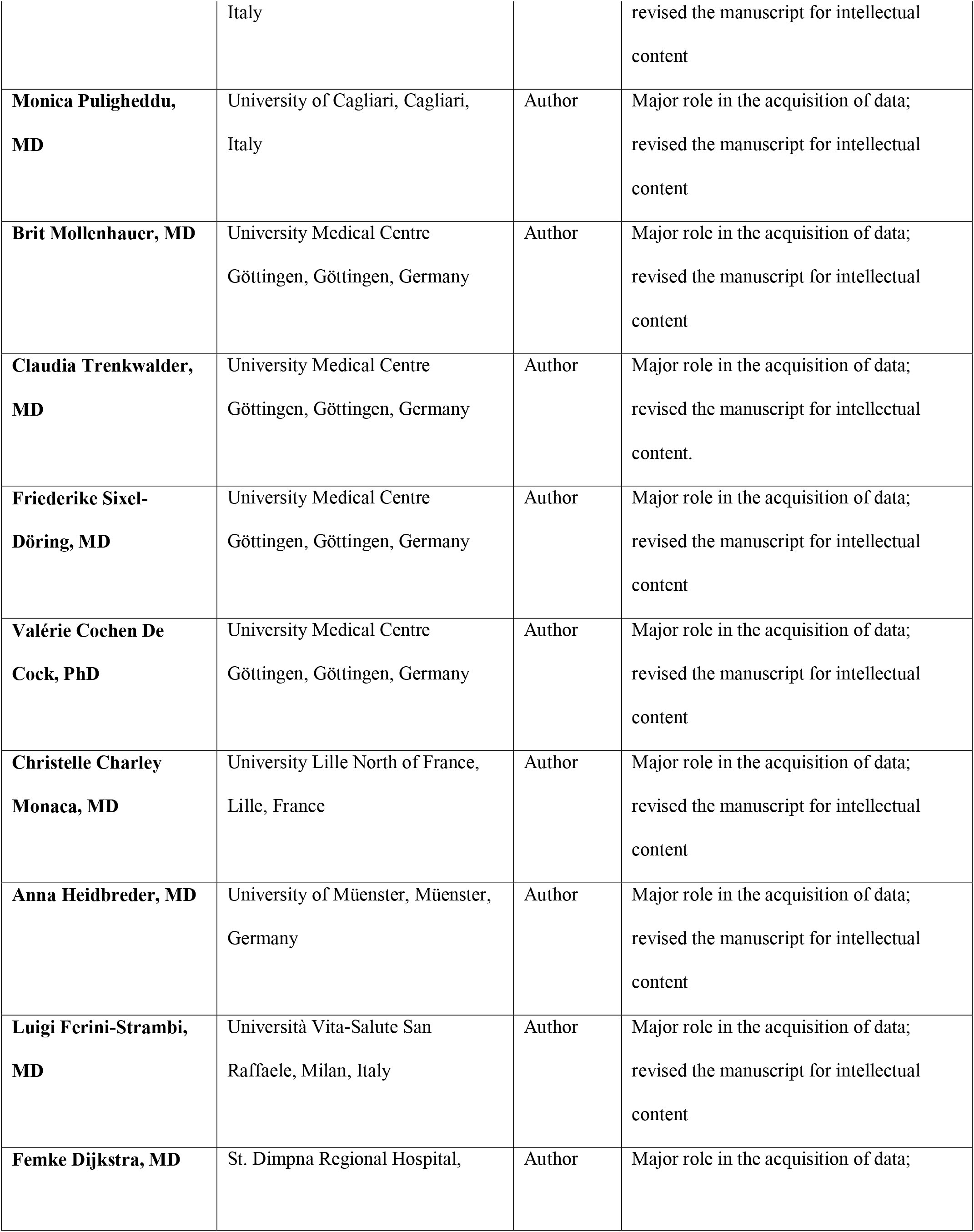
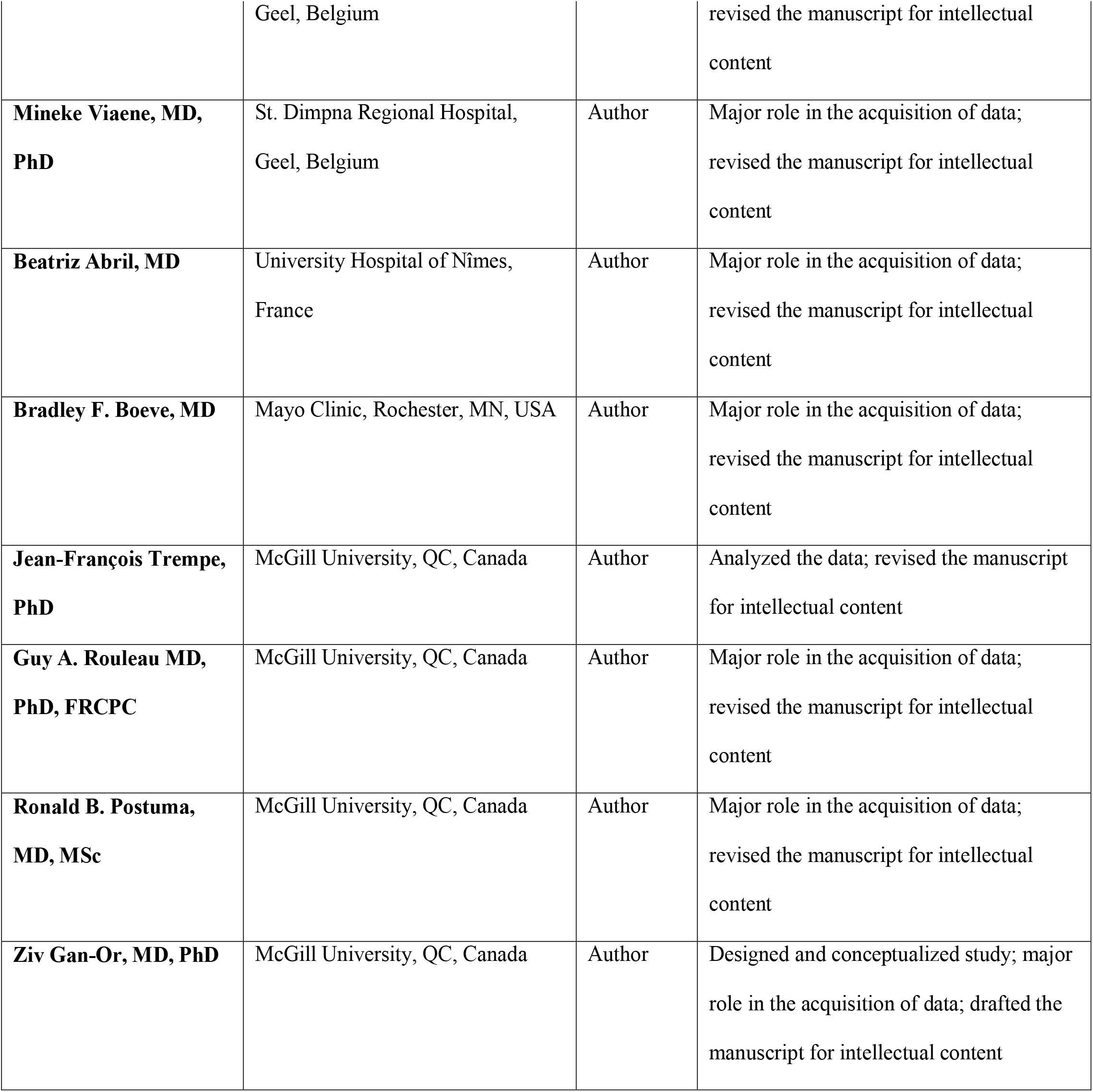

