## Supplemental Table 1 for "Novel associations of *BST1* and *LAMP3* with rapid eye movement sleep behavior disorder"

**Supplementary Table 1. Details on age and sex of samples**

|  | **Number of Samples** | **Sex** | | **Age at sampling (AAS)** | | **Age at onset (AAO)** | | **Age at diagnosis (AAD)** | | **Pheno-conversion to overt neurodegenerative disease** | |
| --- | --- | --- | --- | --- | --- | --- | --- | --- | --- | --- | --- |
|  |  | **Data available** | **Percentage of males** | **Data available** | **Mean** | **Data available** | **Mean** | **Data Available** | **Mean** | **Data available** | **Converted** |
| **iRBD Patients** | 1039 | 1032 | 81% | 1005 | 67.9 ± 9.1 years | 601 | 60.1 ± 10.5 years | 608 | 65.3 ± 8.7 years | 540 | 190 |
| **Controls** | 1852 | 1852 | 51% | 1843 | 52.5 ± 14.3 years | NA | NA | NA | NA | NA | NA |

Abbreviations: iRBD: isolated REM sleep behavior disorder; AAS: age at sampling; AAO: age at onset; AAD: age at diagnosis; NA: non-applicable
