## Supplemental Table 3 for "Novel associations of *BST1* and *LAMP3* with rapid eye movement sleep behavior disorder"

**Supplementary Table 3: Average coverage details for target genes**

| **Gene** | **Average coverage** | **Percent_15x** | **Percent_30x** | **Percent_50x** |
| --- | --- | --- | --- | --- |
| ***SYT11*** | 988 | 100 | 100 | 100 |
| ***RAB25*** | 383 | 100 | 100 | 100 |
| ***RAB7L1*** | 901 | 97 | 97 | 97 |
| ***SLC41A1*** | 809 | 93 | 93 | 93 |
| ***PM20D1*** | 911 | 100 | 100 | 100 |
| ***SIPA1L2*** | 1064 | 100 | 98 | 98 |
| ***TMEM163*** | 891 | 88 | 88 | 88 |
| ***ACMSD*** | 790 | 100 | 100 | 100 |
| ***STK39*** | 883 | 92 | 92 | 92 |
| ***MCCC1*** | 691 | 100 | 100 | 100 |
| ***LAMP3*** | 1162 | 100 | 100 | 97 |
| ***GAK*** | 184 | 98 | 98 | 89 |
| ***DGKQ*** | 73 | 79 | 68 | 54 |
| ***BST1*** | 883 | 100 | 100 | 96 |
| ***GPNMB*** | 1044 | 100 | 100 | 100 |
| ***FGF20*** | 291 | 100 | 100 | 100 |
| ***ITGA8*** | 682 | 98 | 98 | 95 |
| ***CCDC62*** | 911 | 97 | 97 | 97 |
| ***HIP1R*** | 86 | 93 | 82 | 65 |
| ***SETD1A*** | 175 | 89 | 83 | 71 |
| ***STX1B*** | 107 | 88 | 79 | 72 |
| ***MAPT*** | 377 | 96 | 95 | 90 |
| ***RIT2*** | 886 | 100 | 100 | 100 |
| ***DDRGK1*** | 379 | 86 | 86 | 86 |
| ***USP25*** | 627 | 91 | 88 | 83 |
