## Supplemental Table 6 for "Novel associations of *BST1* and *LAMP3* with rapid eye movement sleep behavior disorder"

**Supplementary Table 6. Burden and SKAT-O tests results after excluding the association-driving variants in *BST1* and *LAMP3***

| **DOC** | **Gene** | **All rare (*p* value)** | | **Rare functional (*p* value)** | | **Rare LOF (*p* value)** | | **Rare NS (*p* value)** | | **Rare CADD (*p* value)** | |
| --- | --- | --- | --- | --- | --- | --- | --- | --- | --- | --- | --- |
|  |  | **SKAT-O** | **SKAT Burden** | **SKAT-O** | **SKAT Burden** | **SKAT-O** | **SKAT Burden** | **SKAT-O** | **SKAT Burden** | **SKAT-O** | **SKAT Burden** |
| **30x** | *BST1* | 0.382 | 0.233 | 0.048 | 0.025 | 0.348 | 0.653 | 0.076 | 0.052 | 0.012 | 0.011 |
|  | *LAMP3* | 0.046 | 0.256 | 0.688 | 0.896 | 0.787 | 0.300 | 0.318 | 0.724 | 0.524 | 0.601 |
| **50x** | *BST1* | 0.225 | 0.142 | 0.032 | 0.016 | 0.671 | 0.470 | 0.080 | 0.057 | 0.017 | 0.012 |
|  | *LAMP3* | 0.003 | 0.058 | 0.468 | 0.431 | 0.785 | 0.304 | 0.020 | 0.016 | 0.458 | 0.513 |

DOC: Depth of coverage; LOF: Loss of function; NS: Nonsynonymous; CADD: Combined annotation dependent depletion; SKAT-O: Optimized sequence kernel association test; SKAT: Kernel association test; NV: No variants were found for this filter.
