## Supplemental Table 7 for "Novel associations of *BST1* and *LAMP3* with rapid eye movement sleep behavior disorder"

**Table e-7.** SKAT and SKAT-O analyses of 350 iRBD patients with available data on conversion who did not convert at the time of the study.

| **DOC** | **Gene** | **All rare (*p*-value)** | | **Rare functional (*p*-value)** | | **Rare LOF (*p*-value)** | | **Rare NS (*p*-value)** | | **Rare CADD (*p*-value)** | |
| --- | --- | --- | --- | --- | --- | --- | --- | --- | --- | --- | --- |
|  |  | **SKAT-O** | **SKAT Burden** | **SKAT-O** | **SKAT Burden** | **SKAT-O** | **SKAT Burden** | **SKAT-O** | **SKAT Burden** | **SKAT-O** | **SKAT Burden** |
| **30x** | ***ACMSD*** | 0.649 | 0.398 | 0.712 | 0.654 | NV | NV | 0.712 | 0.654 | 0.712 | 0.654 |
|  | ***BSTI*** | 0.193 | 0.112 | 0.046 | 0.021 | 0.574 | 0.675 | 0.076 | 0.049 | 0.073 | 0.048 |
|  | ***CCDC62*** | 0.034 | 0.519 | 0.650 | 0.473 | 0.573 | 0.676 | 0.756 | 0.815 | 0.588 | 0.953 |
|  | ***DDRGK1*** | 0.010 | 0.030 | 0.199 | 0.273 | 0.224 | 0.162 | 0.745 | 0.951 | 0.232 | 0.414 |
|  | ***DGKQ*** | 0.079 | 0.027 | 0.079 | 0.027 | NV | NV | 0.079 | 0.027 | NV | NV |
|  | ***FGF20*** | 0.050 | 0.062 | 0.214 | 1 | 1 | 1 | 1 | 1 | 1 | 1 |
|  | ***GAK*** | 0.238 | 0.215 | 0.195 | 0.307 | 0.217 | 0.641 | 0.142 | 0.154 | 0.256 | 0.206 |
|  | ***GPNMB*** | 0.128 | 0.716 | 0.217 | 0.153 | 0.109 | 0.836 | 0.524 | 0.395 | 0.582 | 0.634 |
|  | ***HIP1R*** | 0.510 | 0.358 | 0.612 | 1 | NV | NV | 0.283 | 0.150 | 0.285 | 0.172 |
|  | ***ITGA8*** | 0.346 | 0.215 | 0.252 | 0.643 | 0.147 | 0.160 | 0.271 | 0.509 | 0.341 | 0.873 |
|  | ***LAMP3*** | 0.062 | 0.471 | 0.144 | 0.083 | 0.637 | 0.554 | 0.477 | 0.305 | 0.299 | 0.236 |
|  | ***MAPT*** | 0.286 | 0.356 | 0.010 | 0.004 | 1 | 1 | 0.209 | 0.166 | 0.586 | 0.326 |
|  | ***MCCC1*** | 0.122 | 0.064 | 0.154 | 0.092 | 0.148 | 0.162 | 0.071 | 0.045 | 0.061 | 0.038 |
|  | ***PM20D1*** | 0.247 | 0.725 | 0.347 | 0.237 | 0.574 | 0.676 | 0.419 | 0.322 | 0.444 | 0.350 |
|  | ***RAB25*** | 0.383 | 0.253 | 0.647 | 1 | 0.637 | 0.457 | 0.133 | 0.142 | NV | NV |
|  | ***RAB29*** | 0.220 | 0.133 | 0.816 | 0.968 | NV | NV | 0.635 | 0.557 | 0.635 | 0.557 |
|  | ***RIT2*** | 0.431 | 0.317 | 0.135 | 0.340 | NV | NV | 0.278 | 0.743 | 0.278 | 0.743 |
|  | ***SETD1A*** | 0.412 | 0.243 | 0.160 | 0.117 | 0.072 | 0.025 | 0.961 | 1 | 0.837 | 1 |
|  | ***SLC41A1*** | 0.176 | 0.412 | 0.065 | 0.066 | 0.074 | 0.016 | 0.692 | 0.468 | 0.692 | 0.468 |
|  | ***STK39*** | 0.375 | 0.572 | 0.085 | 0.023 | 0.074 | 0.016 | 0.574 | 0.675 | 0.574 | 0.675 |
|  | ***SIPA1L2*** | 0.206 | 0.759 | 0.292 | 0.322 | 0.574 | 0.688 | 0.336 | 0.721 | 0.278 | 0.179 |
|  | ***STX1B*** | 0.692 | 0.464 | NV | NV | NV | NV | NV | NV | NV | NV |
|  | ***SYT11*** | 0.282 | 0.656 | 0.497 | 0.526 | 1 | 1 | 0.574 | 0.675 | 0.574 | 0.675 |
|  | ***TMEM163*** | 0.619 | 0.863 | 0.071 | 0.014 | NV | NV | 0.071 | 0.014 | 0.071 | 0.014 |
|  | ***USP25*** | 0.209 | 0.660 | 0.209 | 0.228 | NV | NV | 0.175 | 0.333 | 0.142 | 0.559 |
| **50x** | ***ACMSD*** | 0.341 | 0.246 | 0.470 | 0.307 | NV | NV | 0.470 | 0.307 | 0.470 | 0.307 |
|  | ***BSTI*** | 0.386 | 0.256 | 0.055 | 0.026 | 0.572 | 0.680 | 0.094 | 0.054 | 0.075 | 0.052 |
|  | ***CCDC62*** | 0.007 | 0.049 | 0.648 | 1 | 1 | 1 | 0.492 | 0.504 | 0.464 | 0.378 |
|  | ***DDRGK1*** | 0.621 | 0.252 | 0.693 | 0.360 | NV | NV | 0.693 | 0.360 | 0.693 | 0.360 |
|  | ***DGKQ*** | NV | NV | NV | NV | NV | NV | NV | NV | NV | NV |
|  | ***FGF20*** | 0.050 | 0.036 | 1 | 1 | 1 | 1 | NV | NV | 1 | 1 |
|  | ***GAK*** | 0.072 | 0.025 | NV | NV | NV | NV | NV | NV | NV | NV |
|  | ***GPNMB*** | 0.058 | 0.500 | 0.791 | 0.609 | 0.091 | 1 | 0.540 | 0.380 | 0.687 | 1 |
|  | ***HIP1R*** | NV | NV | NV | NV | NV | NV | NV | NV | NV | NV |
|  | ***ITGA8*** | 0.242 | 0.763 | 0.280 | 0.634 | 0.143 | 0.150 | 0.355 | 0.790 | 0.343 | 0.653 |
|  | ***LAMP3*** | 0.048 | 1 | 0.484 | 0.204 | 0.633 | 0.458 | 1 | 1 | 0.633 | 0.458 |
|  | ***MAPT*** | 0.707 | 1 | 0.801 | 1 | NV | NV | 0.801 | 1 | 0.801 | 1 |
|  | ***MCCC1*** | 0.045 | 0.021 | 0.272 | 0.120 | 0.136 | 0.122 | 0.124 | 0.066 | 0.110 | 0.058 |
|  | ***PM20D1*** | 0.589 | 1 | 0.444 | 0.280 | 0.567 | 0.584 | 0.555 | 0.334 | 0.584 | 0.361 |
|  | ***RAB25*** | 0.072 | 0.024 | NV | NV | NV | NV | NV | NV | NV | NV |
|  | ***RAB29*** | NV | NV | NV | NV | NV | NV | NV | NV | NV | NV |
|  | ***RIT2*** | 0.892 | 1 | 0.569 | 0.585 | NV | NV | 0.569 | 0.585 | 0.569 | 0.585 |
|  | ***SETD1A*** | 0.460 | 0.282 | 0.572 | 0.596 | NV | NV | 0.572 | 0.596 | 0.572 | 0.596 |
|  | ***SLC41A1*** | 0.324 | 0.501 | NV | NV | NV | NV | NV | NV | NV | NV |
|  | ***STK39*** | 0.096 | 0.138 | 0.068 | 0.023 | 0.068 | 0.023 | 1 | 1 | 1 | 1 |
|  | ***SIPA1L2*** | 0.156 | 0.250 | 0.156 | 0.090 | NV | NV | 0.347 | 0.271 | 0.096 | 0.055 |
|  | ***STX1B*** | NV | NV | NV | NV | NV | NV | NV | NV | NV | NV |
|  | ***SYT11*** | 0.092 | 0.861 | 0.572 | 0.679 | NV | NV | NV | NV | NV | NV |
|  | ***TMEM163*** | 0.445 | 0.562 | 0.070 | 0.023 | NV | NV | 0.070 | 0.023 | 0.070 | 0.023 |
|  | ***USP25*** | 0.337 | 0.274 | 0.111 | 0.109 | NV | NV | 0.111 | 0.109 | 0.471 | 0.401 |

DOC: Depth of coverage; LOF: Loss of function; NS: Nonsynonymous; CADD: Combined annotation dependent depletion; SKAT-O: Optimized sequence kernel association test; SKAT: Kernel association test; NV: No variants were found for this filter.
